# Role of ablation therapy in conjunction with surgical resection for neuroendocrine tumors (NETs): Risks and benefits of multimodality surgical treatment for NETs involving liver

**DOI:** 10.1101/2023.11.01.23297738

**Authors:** Alexander Ostapenko, Stephanie Stroever, Lud Eyasu, Minha Kim, Krist Aploks, Xiang Dong, Ramanathan Seshadri

## Abstract

**Background:** Resection of hepatic metastasis from neuroendocrine tumors (NETs) improves quality of life and prolongs 5-year survival. Ablation can be utilized with surgery to achieve complete resection. Although several studies report long-term outcomes for patients undergoing ablation, none have explored perioperative effects of ablation in patients with metastatic NETs. Our goal was to determine if intra-operative ablation during hepatectomy increases risk of adverse outcomes such as surgical site infections (SSIs), bleeding, and bile leak.

**Methods:** A retrospective analysis of the hepatectomy NSQIP database from 2015-2019 was performed to determine the odds of SSIs, bile leaks, or bleeding in patients undergoing intraoperative ablation when compared to hepatectomy alone.

**Results:** Of the 966 patients included in the study, 298(30.9%) underwent ablation during hepatectomy. There were 78(11.7%) patients with SSIs in the hepatectomy alone group and 39(13.1%) patients with a SSIs in the hepatectomy with ablation group. Bile leak occurred in 41(6.2%) and 14(4.8%) patients in the two groups, respectively; bleeding occurred in 117(17.5%) and 33(11.1%), respectively. After controlling for confounding variables, ablation did not increase risk of SSI (p=0.63), bile leak (p=0.34) or bleeding (p=0.07) when compared to patients undergoing resection alone on multivariate analysis.

**Conclusions:** Intraoperative ablation with hepatic resection for NETs is safe in the perioperative period without significant increased risk of infection, bleeding, or bile leak. Surgeons should utilize this modality when appropriate to achieve optimal disease control and outcomes.

## 1. Introduction

Neuroendocrine tumors (NETs) are epithelial tumors that can arise from most organs. They are indolent, slow growing neoplasms that are frequently discovered at a late stage when they become symptomatic from hormonal excretion by metastasizing to the liver. It is estimated that almost 80-90% of these tumors are inoperable at the time of presentation [1]. However, several studies have demonstrated that resection of hepatic metastasis from NETs improves both quality of life and prolongs 5-year survival [2,3]. These studies demonstrated that aggressive management of hepatic neuroendocrine metastases with adjunct modalities such as transarterial embolization, chemoembolization, and thermal ablation significantly prolong long-term survival and improve patient outcomes [2,4]. Despite these findings there is hesitancy among surgeons to utilize radiofrequency ablation due to fear of bleeding, abdominal infections, and biliary tree injury [5,6].

Of the available image guided ablative therapies, ethanol, microwave, radiofrequency, and cyro-ablation are the most common. These adjuncts have been utilized for primary malignancies such as hepatocellular carcinoma and cholangiocarcinoma and are now accepted as the curative treatment option for early HCC by most guidelines [4, 35]. Similarly ablation has been utilized for metastatic disease for colorectal, breast, and NETs [1,4,36,38]. There is a scarcity of literature and guidelines on ablation for metastatic NETs, likely in part due to the rarity of the disease when compared to other liver metastasis. Several authors have reported their individual experience with ablation with NETs, claiming that it is a safe and efficacious treatment modality, but no multi-center studies or national database studies have been reported[16]. In this study, we set out to explore the effects of intra-operative ablation in patients undergoing hepatic resection for NETs. Using the American College of Surgeons (ACS) National Surgery Quality Improvement Program (NSQIP) database and Hepatectomy Procedure Targeted database we compared the rates of adverse outcomes in patients undergoing surgery alone versus surgery with intraoperative ablation. We hypothesize that patients who had liver ablation concurrently with their hepatectomy did not have significantly higher rates of bleeding, surgical site infections, bile leaks, and readmission.

## 2. Materials and Methods

### 2.1 The Study Design and Participants

We performed a cross sectional study utilizing ACS-NSQIP database and Hepatectomy Procedure Targeted database for 2015-2019. The variables from the Procedure Targeted database were merged with the standard public use file (PUF) using the CASEID variable. We included patients undergoing hepatic resection for neuroendocrine liver metastasis who were between the age of 18 and 90. Patients were separated into two study groups: those who underwent an intra-operative ablation concurrently with a hepatectomy and those undergoing hepatectomy alone.

### 2.2 Variables

The aim of this study was to determine if intra-operative ablation increases the risk of adverse outcomes in patients undergoing hepatic resection for neuroendocrine liver metastasis. We included microwave, RFA, and alcohol ablation. The adverse outcomes of interest included surgical site infections, bleeding, bile leak, and readmission. We evaluated for the risk factors predictive of the adverse outcomes that were significantly different between the groups as possible confounders which could contribute to the outcomes of interest.

### 2.3 Statistical Analysis

StataSE was used for statistical analysis. Descriptive statistics including mean/SD for normally distributed continuous variables, median/interquartile range for skewed continuous variables, and number/percentage for categorical variables. We assessed univariate differences in outcomes between patients who underwent ablation concurrently with resection and those with resection alone using the chi-square test for categorical variables. Variables that were statistically associated (α < 0.05) with both the outcome and ablation were included in multivariate logistic regression analysis. The multivariable logistic regression analysis was performed to determine if the risk of developing adverse outcomes was higher among patients undergoing ablation, while controlling for confounding covariates. We used stepwise, backward selection and tested full/reduced models with the likelihood ratio test to determine the most parsimonious model. A p-value >0.05 indicated that the reduced model fit as well as the full model and the removed variable was unnecessary. A p-value < 0.05 indicated that the full model was better, and the removed variable should be maintained.

## 3. Results

### 3.1. Patient and dataset characteristics

The patients were categorized by gender, age, race, ethnicity, comorbidities (BMI, diabetes), pre-operative factors such as steroid use, albumin, and neoadjuvant chemotherapy, wound class assigned to the surgery, and specific tumor characteristics (Table 1). There were 966 patients included in the study. There were 298 patients (30.8%) who had intraoperative ablation concurrently with hepatic resection and 688(69.2%) had hepatic resection alone.

**Table 1:**
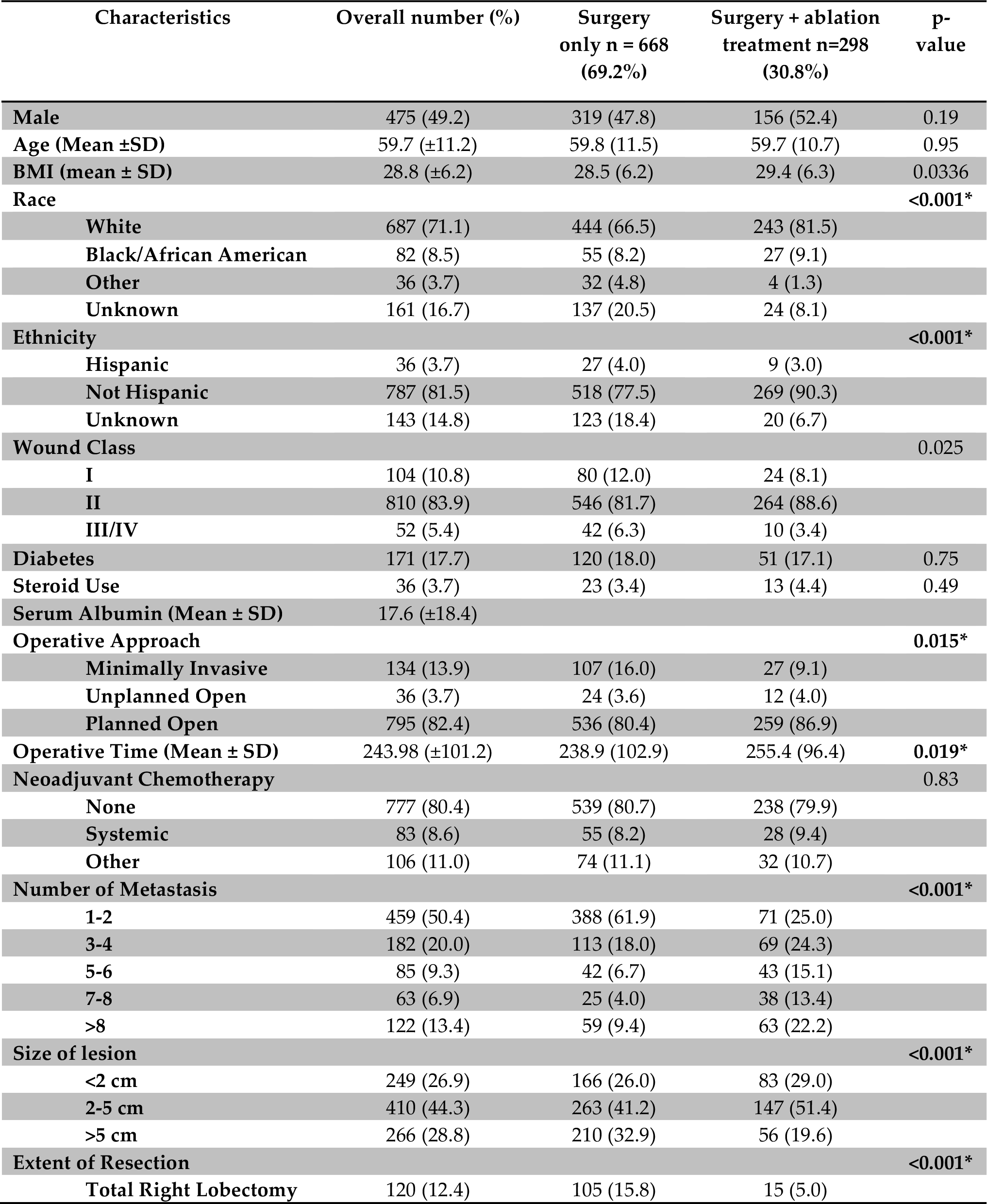
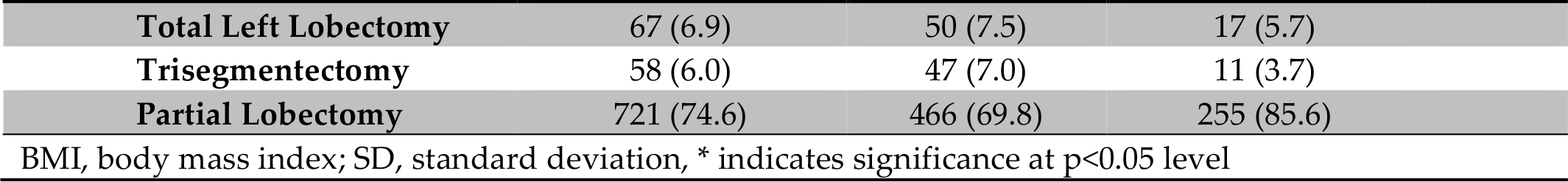
Demographic, clinical characteristics, and descriptive statistics of patients who underwent resection for metastatic neuroendocrine tumors to the liver who were included in the study and were used to assess determinants of adverse outcomes(n=966).

| Characteristics | Overall number (%) | Surgery only n = 668 (69.2%) | Surgery + ablation treatment n=298 (30.8%) | p-value |
| --- | --- | --- | --- | --- |
| Male | 475 (49.2) | 319 (47.8) | 156 (52.4) | 0.19 |
| Age (Mean ±SD) | 59.7 (±11.2) | 59.8 (11.5) | 59.7 (10.7) | 0.95 |
| BMI (mean ± SD) | 28.8 (±6.2) | 28.5 (6.2) | 29.4 (6.3) | 0.0336 |
| Race |  |  |  | <0.001* |
| White | 687 (71.1) | 444 (66.5) | 243 (81.5) |  |
| Black/African American | 82 (8.5) | 55 (8.2) | 27 (9.1) |  |
| Other | 36 (3.7) | 32 (4.8) | 4 (1.3) |  |
| Unknown | 161 (16.7) | 137 (20.5) | 24 (8.1) |  |
| Ethnicity |  |  |  | <0.001* |
| Hispanic | 36 (3.7) | 27 (4.0) | 9 (3.0) |  |
| Not Hispanic | 787 (81.5) | 518 (77.5) | 269 (90.3) |  |
| Unknown | 143 (14.8) | 123 (18.4) | 20 (6.7) |  |
| Wound Class |  |  |  | 0.025 |
| I | 104 (10.8) | 80 (12.0) | 24 (8.1) |  |
| II | 810 (83.9) | 546 (81.7) | 264 (88.6) |  |
| III/IV | 52 (5.4) | 42 (6.3) | 10 (3.4) |  |
| Diabetes | 171 (17.7) | 120 (18.0) | 51 (17.1) | 0.75 |
| Steroid Use | 36 (3.7) | 23 (3.4) | 13 (4.4) | 0.49 |
| Serum Albumin (Mean ± SD) | 17.6 (±18.4) |  |  |  |
| Operative Approach |  |  |  | 0.015* |
| Minimally Invasive | 134 (13.9) | 107 (16.0) | 27 (9.1) |  |
| Unplanned Open | 36 (3.7) | 24 (3.6) | 12 (4.0) |  |
| Planned Open | 795 (82.4) | 536 (80.4) | 259 (86.9) |  |
| Operative Time (Mean ± SD) | 243.98 (±101.2) | 238.9 (102.9) | 255.4 (96.4) | 0.019* |
| Neoadjuvant Chemotherapy |  |  |  | 0.83 |
| None | 777 (80.4) | 539 (80.7) | 238 (79.9) |  |
| Systemic | 83 (8.6) | 55 (8.2) | 28 (9.4) |  |
| Other | 106 (11.0) | 74 (11.1) | 32 (10.7) |  |
| Number of Metastasis |  |  |  | <0.001* |
| 1-2 | 459 (50.4) | 388 (61.9) | 71 (25.0) |  |
| 3-4 | 182 (20.0) | 113 (18.0) | 69 (24.3) |  |
| 5-6 | 85 (9.3) | 42 (6.7) | 43 (15.1) |  |
| 7-8 | 63 (6.9) | 25 (4.0) | 38 (13.4) |  |
| >8 | 122 (13.4) | 59 (9.4) | 63 (22.2) |  |
| Size of lesion |  |  |  | <0.001* |
| <2 cm | 249 (26.9) | 166 (26.0) | 83 (29.0) |  |
| 2-5 cm | 410 (44.3) | 263 (41.2) | 147 (51.4) |  |
| >5 cm | 266 (28.8) | 210 (32.9) | 56 (19.6) |  |
| Extent of Resection |  |  |  | <0.001* |
| Total Right Lobectomy | 120 (12.4) | 105 (15.8) | 15 (5.0) |  |

|  |  |  |  |
| --- | --- | --- | --- |
| **Total Left Lobectomy** | 67 (6.9) | 50 (7.5) | 17 (5.7) |
| **Trisegmentectomy** | 58 (6.0) | 47 (7.0) | 11 (3.7) |
| **Partial Lobectomy** | 721 (74.6) | 466 (69.8) | 255 (85.6) |

BMI, body mass index; SD, standard deviation, \* indicates significance at p<0.05 level

### 3.2. Outcomes

The rates of adverse outcomes are listed in Table 2. There were no significant differences in most adverse outcomes between patients who underwent intra-operative ablation versus hepatectomy alone on univariate regression analysis. However, there was a significant difference between bleeding (p=0.011) with higher rates of bleeding seen in the hepatectomy only group (Table 2).

**Table 2:** Outcomes of patients treated for metastatic neuroendocrine tumors to the liver (n=966)

| **Outcome** | **Surgical treatment**<br>**only n = 668 (69.2%)** | **Surgery + ablation**<br>**treatment n=298 (30.8%)** | **P-value** |
| --- | --- | --- | --- |
| **Death** | 7 (1.1) | 3 (1.0) | 1.00 |
| **Significant Bleed** | 117 (17.5) | 33 (11.1) | **0.011\*** |
| **Bile Leak** | 41 (6.2) | 14 (4.8) | 0.38 |
| **Myocardial Infarction** | 3 (0.5) | 1 (0.3) | 1.00 |
| **Pulmonary Embolism** | 11 (1.7) | 4 (1.3) | 1.00 |
| **Pneumonia** | 17 (2.5) | 8 (2.7) | 0.90 |
| **Sepsis** | 21 (3.1) | 16 (5.4) | 0.096 |
| **Liver Failure** | 23 (3.4) | 4 (1.3) | 0.089 |
| **Return to Operating Room** | 24 (3.6) | 7 (2.4) | 0.31 |
| **Readmission** | 73 (10.9) | 34 (11.4) | 0.83 |
| **Surgical Site infection** |  |  |  |
| **Superficial** | 27 (4.0) | 10 (3.4) | 0.61 |
| **Deep incisional** | 4 (0.6) | 1 (0.3) | 1.00 |
| **Organ space** | 48 (7.2) | 30 (10.1) | 0.13 |
| **Wound** | 4 (0.6) | 1 (0.3) | 1.00 |
| **Any** | 78 (11.7) | 39 (13.1) | 0.54 |

BMI, body mass index; SD, standard deviation; \* indicates significance at p<0.05 level

### 3.3. Univariate Analysis

We performed a univariate analysis of variables predictive of bile leaks, readmission, bleeding, organ space surgical site infections, and any surgical infection. There were 55(5.7%) patients who experienced a bile leak. None of the variables in our model, included intra-operative ablation, were predictive of a bile leak occurrence (Supplemental table 1). There were 107(11.1%) patients who required a readmission within 30 days of surgery. Similarly, none of the variables including ablation were predictive of readmission (Supplemental Table 2).

There were 150(15.5%) significant bleeding occurrences that required blood transfusion (Supplemental Table 3). We determined several factors predictive of bleeding. A minimally invasive surgical approach (OR=0.45, p=0.015), partial lobectomy (OR=0.50, p=0.005), and ablation (OR=0.59, p=0.011) were protective from bleeding. On the other hand, neoadjuvant chemotherapy (OR=2.55, p<0.001), wound class II (OR=3.38, p=0.006), and lesion size between 2-5cm (OR=2.56, p=0.005) all increased risk of significant bleeding. Furthermore, patients undergoing hepatecomy for lesions greater than 5cm had 8.05 times the odds of bleeding than patients with lesions smaller than 2cm (OR=8.05, p<0.001). However, extensive hepatectomy such as trisegmentectomy or total left hepatectomy did not confer increased risk of bleeding.

There were insufficient occurrences of superficial, deep, and wound dehiscence for regression modeling. We therefore only performed univariate analysis for organ space infections and any surgical site infections. There were 78(8.1%) organ space infections (Supplemental Table 4). Most of the variables included in the model were not predictive of infections; however Hispanic ethnicity (OR=3.97, p<0.001) and presence of more than 8 metastases (OR=2.16, p=0.013) increased the risk of organ space SSI. These findings were similar for any surgical site infection (Supplemental Table 5). We therefore performed a multivariable analysis controlling for significant covariates to determine if intra-operative ablation increased risk of the adverse outcomes of interest.

### 3.4 Multivariate Analysis

On multivariate logistic regression analysis, there were no differences in adverse outcomes between patients undergoing intraoperative ablation and those undergoing hepatectomy alone (Figure 3). After controlling for potential confounding covariates that were identified in the univariate analysis, we determined the odds of readmission (OR 0.88, p=0.64), bile leaks (OR 0.69, p=0.34), organ space SSIs (OR 1.10, p=0.74), or any SSIs (OR 0.89, p=0.63), which were not significantly different between the two groups. Although ablation was associated with significant bleeding on univariate analysis, after controlling for the predictive covariates in the multivariate analysis it was no longer predictive of bleeding (OR 0.61, p=0.07).

**Figure 3:**
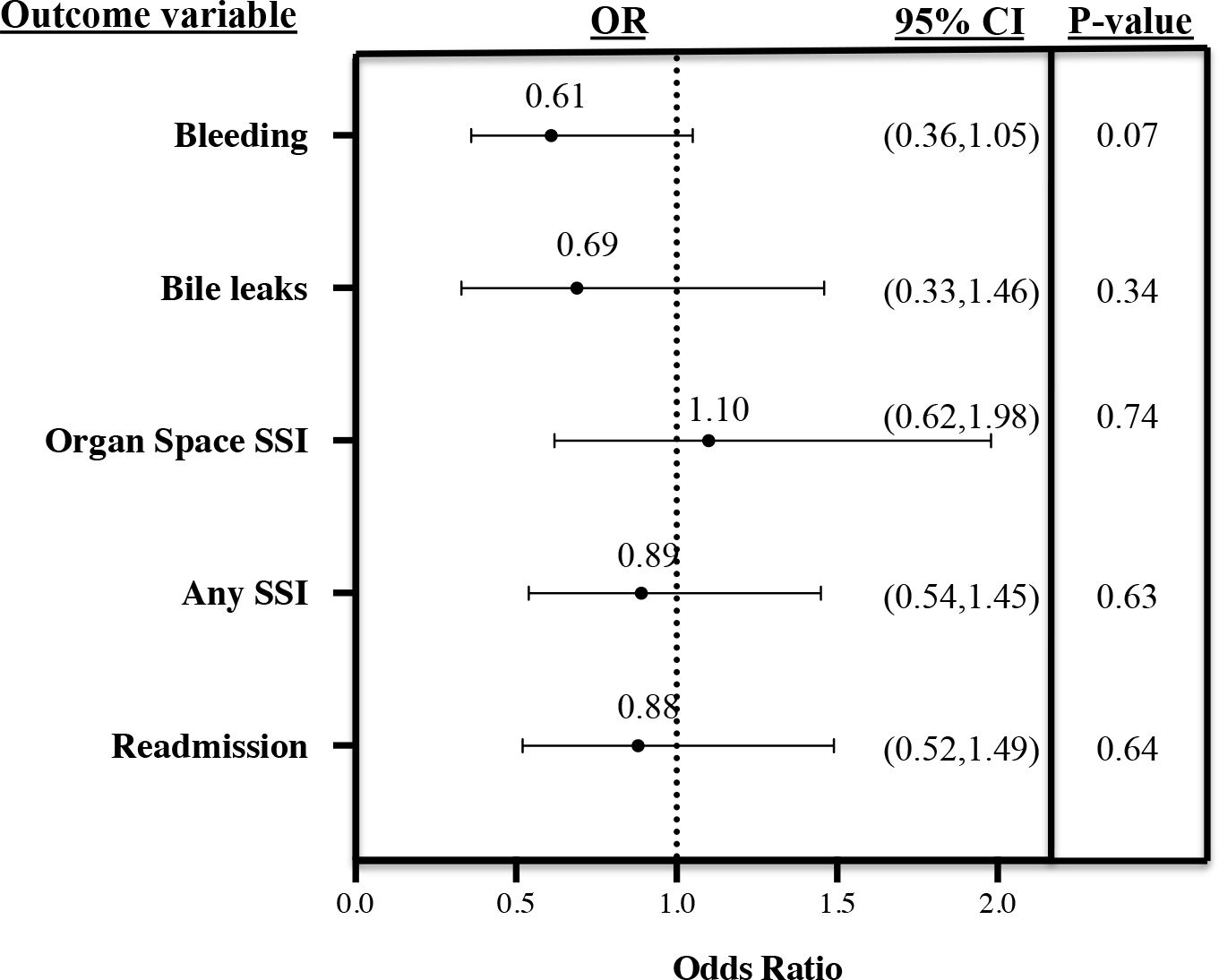
Forest plot depiction of the multivariable regression model for predictors of outcomes among patients treated with concurrent intraoperative ablation compared to surgery alone. OR, odds ratio; CI, confidence interval; SSI, surgical site infection.

## 4. Discussion

Management of liver malignancies is complex and is primarily driven by the pathology, size, location, and number of tumors. The surgical approach to liver malignancies has changed dramatically over the last few decades with the introduction and utilization of new technology, which has facilitated operative planning and improved surgical precision [7]. This has resulted in a marked decline in many perioperative complications [8].

Several adjunct modalities to surgery have been utilized to improve surgical outcomes, including portal vein embolization, stereotactic body radiation therapy, and thermal/chemical ablation. Ablation utilization has been rising over time for both primary liver and biliary tumors as well as metastatic colorectal disease [9-11]. The reported incidence of significant complications from ablation modalities ranges between 6-11%[12,13]. Most studies report bleeding, abdominal infection, biliary track damage, portal vein thrombosis, and pleural effusion/pneumothorax as the most frequent complications, causing hesitancy in widespread utilization of ablation [5,12,14]. Despite this, the use of ablation therapy has been applied to patients with a variety of disease pathologies, including metastatic neuroendocrine tumors to the liver.

Treatment algorithms for hepatic NETs metastases have changed over time. Non-operative management with chemotherapy, external beam radiation, and octreotide has been shown to be significantly inferior to surgical management [2]. Touzios et al. demonstrated that patients who did not undergo surgery had a median survival of 20 months while patients who underwent resection with or without ablation had a median survival of greater than 96 months. Furthermore, an aggressive approach to these lesions not only improved survival, but also provided symptomatic relief from the release of active hormones. Up to 95% of patients in the operative group experienced symptom relief, while only 42% in the non-operative group had similar response. Therefore, aggressive surgical approach with adjuncts such as radiofrequency ablation and trans-arterial chemoembolization (TACE) has grown in popularity for both symptom control and survival benefits^4^. Prior literature has established that curative intent is the best treatment option for hepatic metastasis from NETs.

Given the evidence for aggressive surgical control of metastatic NETs, we set out to explore the perioperative risks of ablation in conjunction with hepatic resection. Several studies report the overall morbidity of ablation used concurrently with resection to be between 20 and 42% [2,15]. However, these studies were limited by insufficient sample sizes for multivariate analysis as well as use of composite outcomes; as a result, it is difficult to determine whether there is an association between ablation therapy and specific adverse outcomes. Studies describing ablation monotherapy in patients with unresectable NETs hepatic metastasis were similarly restricted by sample size and use of composite outcomes, as well as being limited by lack of generalizability [16,17]. In our study we report the frequencies of each adverse outcome and demonstrate that intra-operative ablation does not increase risk of common complications when compared to hepatectomy alone.

This study represents the largest sample of patients to date with hepatic NET metastases treated simultaneously with ablation and surgery. Of the 966 patients included in the study, 298(30.8%) underwent ablation concurrently with a resection. In principle, ablation is combined with surgical resection in patients with bilateral tumors, for tumors deep in the liver parenchyma, or when resections of many individual lesions would significantly decrease the future liver remnant (FLR) [2]. We therefore expected patients in the ablation group to have large tumors, require extensive hepatectomy, or have greater number of lesions. Although the ablation group had fewer extensive hepatic resections (total lobectomy or trisemgmentectomies), there were more patients with >8 tumors and with greater number of smaller lesions. This was unsurprising, as previous studies reported higher efficacy of ablation for smaller lesions [15,17,18].

The rate of significant bleeding in our study was 15.5%. The reported rates in the literature vary widely between 1.1% and 25%, likely due to the subjective nature of “significant bleeding”[19,20]. Unsurprisingly, minimally invasive approach and smaller resections in the form of partial lobectomy were protective from bleeding. On the other hand, neoadjuvant chemotherapy and larger lesions increased risk of significant bleeding, with the greatest risk in patients with lesions greater than 5cm. When potential confounders were included in the multivariate model, intra-operative ablation did not confer a significant risk on bleeding (OR 0.61, p=0.07) when compared to surgery alone (Table 3). This finding is congruent with current literature of smaller case series [15]. Although there are isolated case reports of massive hemorrhage after ablation, they are unique cases that focus on extraneous factors contributing to the bleeding [21,22].

Biliary complications and bile leaks occur in 3% to 12% of hepatic resections, and tend to confer significant morbidity due to need for percutaneous drainage, endoscopic retrograde cholangiopancreatography, or surgical intervention [23-25]. In our study there were 55(5.7%) bile leaks occurrences, with no significant difference between patients who had ablation and those who underwent hepatectomy alone. Even after controlling for possible predictive covariates in the multivariate analysis, ablation did not significantly increase risk of bile leaks. Capussotti et al. reported specific resections such as a left hepatectomy including segment 1 or 4 as independent predictor of bile leak [23]. However, in our study none of the covariates tested in the multivariate model, including type of resection, neoadjuvant chemotherapy, ablation, or operative approach were predictive of bile leaks.

There were 177(12.1%) patients who developed an SSI in our study, the majority of which were organ space infections (8.1%). In a series of 81 patients undergoing either hepatic resection or ablation, Karavokyros et al. reported organ space infection in 11.1% of patients, although notably the study included synchronous colorectal resection as well [26]. Ramanathan et al. included patients undergoing hepatectomy for cholangiocarcinoma, and reported a rate of 17.6% of organ space infections [27]. Other studies explored hepatic abscesses after percutaneous ablation alone without surgery. In this population the frequency of abscesses is significantly lower, ranging between 1% and 4%[28,29]. Risk factors most predictive of abscess are transarterial chemoembolization and again biliary abnormalities predisposing to ascending infections. In fact, the frequency of liver abscesses after ablation in patients with a history of sphincter of Odi manipulation via sphincterototomy, stenting, or biliary drainage ranges between 20% and 75%[30-32]. In our study we did not include pre-operative stent in our analysis. However, we performed multivariate analysis exploring other covariates that were predictive of organ space or any SSI. We found that Hispanic ethnicity had 3.97 and 2.68 times the odds of developing an organ space or any SSI respectively. Similarly having greater than 8 metastatic lesions increased the odds by 2.16 and 1.84 of an organ space or any SSI respectively. Interestingly a minimally invasive surgical approach was protective against any SSI when compared to open approach (OR 0.36, p=0.012) on univariate analysis, although on multivariate analysis it did not have a significant effect. Intraoperative ablation did not increase risk of either organ space or any SSI on univariate and multivariate analysis.

There are several limitations in this study that should be addressed. This is a cross-sectional retrospective study and is inferior to randomized control trials. Limitations of ACS-NSQIP data that are widely recognized and frequently described apply to this study and include scope of available variables, no mechanism of external data validation, and short-term 30-day outcomes [33-35]. For example, this study could be enhanced if the primary source of the metastatic NET was available in order to delineate pancreatic from carcinoid origin, as well as information about the location or size of the ablated tumor, number of ablated and resected tumors, or involvement of major vascular branches; however, these variables are not reported. Similarly, lacking is the reason surgeons chose to perform intraoperative ablation. Despite these limitations, NSQIP offers a robust database with a large number of patients sampled from across the country, useful for exploring rare pathologies such as metastatic NETs. The database is ideal for a study design exploring perioperative outcomes with specific interventions.

## 4. Conclusion

Liver metastases from neuroendocrine tumors have an indolent course, often remaining undetected until hormonal oversecretion or symptoms from mass effect. Surgical resection remains the best therapy for both survival benefit and symptom control when complete irradiation is achieved. Ablation therapies are efficacious adjuncts to accomplish this goal. In this study we demonstrate that ablation is safe in the perioperative period and does not increase the risk of infection, bleeding, or bile leak in patients undergoing hepatic resection for neuroendocrine liver metastasis. Surgeons should not fear utilizing ablation concurrently with resection, even when attempting complete resection.

## Data Availability

All data produced are available online at NSQIP

https://www.facs.org/quality-programs/data-and-registries/acs-nsqip/participant-use-data-file/participant-use-request-form/

## Institutional Review Board Statement

This study was exempt from our Institutional Review Board review as the data was deidentified and obtained from a public use file.

## Informed Consent Statement

Patient consent was waived due to deidentified patient information obtained from a public use file.

## Data Availability Statement

All data was acquired from public use files obtain from the ACS National Surgical Quality Improvement Program at (www.facs.org/quality-programs/acs-nsqip), following all acquisition requirements including the user agreement contract.

