## Supplemental Table 1 for "Role of ablation therapy in conjunction with surgical resection for neuroendocrine tumors (NETs): Risks and benefits of multimodality surgical treatment for NETs involving liver"

**Supplemental Table 1:** Univariate analysis of patient and procedure-related determinants of **bile leak (n=55, 5.7%)** in patients undergoing resection of metastatic neuroendocrine tumors to the liver.

| Variable | Odds ratio | 95% CI | P value |
| --- | --- | --- | --- |
| Intraoperative ablation | 0.76 | 0.41, 1.41 | 0.38 |
| Age | 0.98 | 0.96, 1.00 | 0.089 |
| Sex | 1.16 | 0.67, 2.01 | 0.59 |
| Race group |  |  |  |
| Black/African American | 0.38 | 0.09, 1.60 | 0.19 |
| Other | 0.90 | 0.21, 3.86 | 0.88 |
| Not reported | 0.91 | 0.44, 1.92 | 0.81 |
| Ethnic group |  |  |  |
| Hispanic | 0.97 | 0.23, 4.17 | 0.97 |
| Not reported | 0.84 | 0.37 1.89 | 0.67 |
| Wound classification |  |  |  |
| I | 2.02 | 0.62, 6.63 | 0.24 |
| II | 4.39 | 1.00, 19.25 | 0.050 |
| III/IV | 4.17 | 0.39, 44.80 | 0.24 |
| BMI | 0.98 | 0.94, 1.03 | 0.41 |
| Obese (BMI>30) | 1.18 | 0.68, 2.06 | 0.56 |
| Diabetes | 0.44 | 0.17, 1.13 | 0.089 |
| Steroid Use | 0.97 | 0.23, 4.13 | 0.96 |
| Operative approach |  |  |  |
| Any MIS | 0.62 | 0.24, 1.58 | 0.31 |
| Unplanned open | 1.44 | 0.42, 4.85 | 0.56 |
| Neoadjuvant chemotherapy |  |  |  |
| Pre op systemic | 1.64 | 0.71, 3.78 | 0.25 |
| All other | 1.26 | 0.55, 2.88 | 0.59 |
| Number of mets resected |  |  |  |
| 3 – 4 | 0.52 | 0.21, 1.28 | 0.16 |
| 5 – 6 | 0.95 | 0.36, 2.54 | 0.92 |
| 7 – 8 | 0.51 | 0.12, 2.19 | 0.36 |
| > 8 | 1.36 | 0.64, 2.89 | 0.42 |
| Size of lesion |  |  |  |
| 2-5 cm | 1.30 | 0.60, 2.81 | 0.50 |
| > 5cm | 2.15 | 1.00, 4.65 | 0.050 |
| Extent of resection |  |  |  |
| Total left lobectomy | 0.89 | 0.26, 3.07 | 0.85 |
| Trisegmentectomy | 1.32 | 0.41, 4.23 | 0.64 |
| Partial lobectomy | 0.79 | 0.36, 1.73 | 0.55 |
| CI, confidence interval; MIS, minimally invasive surgery; * indicates significance at p<0.05 level | | | |
