## Supplemental Table 2 for "Role of ablation therapy in conjunction with surgical resection for neuroendocrine tumors (NETs): Risks and benefits of multimodality surgical treatment for NETs involving liver"

**Supplemental Table 2:** Univariate analysis of patient and procedure-related determinants of **readmission (n=107, 11.7%)** in patients undergoing resection of metastatic neuroendocrine tumors to the liver.

| Variable | Odds ratio | 95% CI | P value |
| --- | --- | --- | --- |
| Intraoperative ablation | 1.05 | 0.68, 1.62 | 0.83 |
| Age | 0.97 | 0.95, 0.98 | **<0.001*** |
| Sex | 1.21 | 0.81, 1.82 | 0.35 |
| Race group |  |  |  |
| Black/African American | 0.80 | 0.37, 1.71 | 0.56 |
| Other | 0.67 | 0.20, 2.24 | 0.52 |
| Not reported | 0.70 | 0.39, 1.27 | 0.25 |
| Ethnic group |  |  |  |
| Hispanic | 1.85 | 0.79, 4.34 | 0.16 |
| Not reported | 0.51 | 0.25, 1.04 | 0.066 |
| Wound classification |  |  |  |
| I | 1.12 | 0.56, 2.22 | 0.75 |
| II | 2.49 | 0.93, 6.64 | 0.069 |
| III/IV | 2.69 | 0.49, 14.72 | 0.26 |
| BMI | 1.01 | 0.97, 1.04 | 0.77 |
| Obese (BMI>30) | 1.42 | 0.94, 2.13 | 0.94 |
| Diabetes | 1.08 | 0.64, 1.81 | 0.78 |
| Steroid Use | 1.00 | 0.35, 2.90 | 1.00 |
| Operative approach |  |  |  |
| Any MIS | 0.86 | 0.47, 1.59 | 0.64 |
| Unplanned open | 1.61 | 0.65, 3.97 | 0.30 |
| Neoadjuvant chemotherapy |  |  |  |
| Pre op systemic | 1.55 | 0.82, 2.93 | 0.17 |
| All other | 0.97 | 0.50, 1.88 | 0.92 |
| Number of mets resected |  |  |  |
| 3 – 4 | 0.76 | 0.43, 1.34 | 0.34 |
| 5 – 6 | 0.76 | 0.35, 1.67 | 0.50 |
| 7 – 8 | 0.63 | 0.24, 1.65 | 0.35 |
| > 8 | 1.03 | 0.56, 1.89 | 0.93 |
| Size of lesion |  |  |  |
| 2-5 cm | 0.89 | 055, 1.44 | 0.63 |
| > 5cm | 0.79 | 0.46, 1.37 | 0.41 |
| Extent of resection |  |  |  |
| Total left lobectomy | 1.74 | 0.70, 4.34 | 0.24 |
| Trisegmentectomy | 1.36 | 0.50, 3.71 | 0.55 |
| Partial lobectomy | 1.22 | 0.63, 2.37 | 0.56 |
| CI, confidence interval; MIS, minimally invasive surgery; * indicates significance at p<0.05 | | | |
