## Supplemental Table 3 for "Role of ablation therapy in conjunction with surgical resection for neuroendocrine tumors (NETs): Risks and benefits of multimodality surgical treatment for NETs involving liver"

**Supplemental Table 3:** Univariate analysis of patient and procedure-related determinants of **significant bleeding (n=150, 15.5%)** in patients undergoing resection of metastatic neuroendocrine tumors to the liver.

| Variable | Odds ratio | 95% CI | P value |
| --- | --- | --- | --- |
| Intraoperative ablation | 0.59 | 0.39, 0.89 | **0.011*** |
| Age | 0.99 | 0.97, 1.00 | 0.14 |
| Sex | 1.41 | 0.99, 2.00 | 0.057 |
| Race group |  |  |  |
| Black/African American | 1.5 | 0.85, 2.66 | 0.17 |
| Other | 1.91 | 0.87, 4.18 | 0.11 |
| Not reported | 0.91 | 0.55, 1.49 | 0.70 |
| Ethnic group |  |  |  |
| Hispanic | 1.30 | 0.56, 3.04 | 0.54 |
| Not reported | 0.88 | 0.53, 1.46 | 0.62 |
| Wound classification |  |  |  |
| I | 1.22 | 0.66, 2.25 | 0.53 |
| II | 3.38 | 1.43, 8.01 | **0.006*** |
| III/IV | 3.5 | 0.78, 15.73 | 0.10 |
| BMI | 0.97 | 0.94, 1.00 | **0.044*** |
| Obese (BMI>30) | 0.76 | 0.52, 1.10 | 0.15 |
| Diabetes | 1.20 | 0.77, 1.86 | 0.42 |
| Steroid Use | 1.59 | 0.71, 3.55 | 0.26 |
| Operative approach |  |  |  |
| Any MIS | 0.45 | 0.24, 0.86 | **0.015*** |
| Unplanned open | 1.21 | 0.52, 2.83 | 0.66 |
| Neoadjuvant chemotherapy |  |  |  |
| Pre op systemic | 2.55 | 1.52, 4.27 | **<0.001*** |
| All other | 1.37 | 0.80, 2.34 | 0.25 |
| Number of mets resected |  |  |  |
| 3 – 4 | 0.78 | 0.46, 1.30 | 0.34 |
| 5 – 6 | 0.59 | 0.27, 1.27 | 0.18 |
| 7 – 8 | 0.94 | 0.44, 2.00 | 0.88 |
| > 8 | 1.53 | 0.93, 2.53 | 0.097 |
| Size of lesion |  |  |  |
| 2-5 cm | 2.56 | 1.33, 4.92 | **0.005*** |
| > 5cm | 8.05 | 4.25, 15.22 | **<0.001*** |
| Extent of resection |  |  |  |
| Total left lobectomy | 0.79 | 0.38, 1.66 | 0.534 |
| Trisegmentectomy | 0.95 | 0.45, 2.01 | 0.89 |
| Partial lobectomy | 0.50 | 0.31, 0.81 | **0.005*** |
| CI, confidence interval; MIS, minimally invasive surgery; * indicates significance at p<0.05 | | | |
