## Supplemental Table 4 for "Role of ablation therapy in conjunction with surgical resection for neuroendocrine tumors (NETs): Risks and benefits of multimodality surgical treatment for NETs involving liver"

**Supplemental Table 4:** Univariate analysis of patient and procedure-related determinants of **organ space surgical site infection (n=78, 8.1%)** in patients undergoing resection of metastatic neuroendocrine tumors to the liver.

| Variable | Odds ratio | 95% CI | P value |
| --- | --- | --- | --- |
| Intraoperative ablation | 1.45 | 0.90, 2.33 | 0.13 |
| Age | 0.99 | 0.97, 1.01 | 0.39 |
| Sex | 0.96 | 0.61, 1.53 | 0.88 |
| Race group |  |  |  |
| Black/African American | 0.57 | 0.20, 1.60 | 0.29 |
| Other | 1.00 | 0.30, 3.38 | 0.99 |
| Not reported | 1.05 | 0.57, 1.94 | 0.87 |
| Ethnic group |  |  |  |
| Hispanic | 3.97 | 1.79, 8.81 | **0.001*** |
| Not reported | 0.71 | 0.33, 1.51 | 0.37 |
| Wound classification |  |  |  |
| I | 1.21 | 0.54, 2.71 | 0.65 |
| II | 1.82 | 0.55, 6.10 | 0.33 |
| III/IV | 1.73 | 0.19, 15.88 | 0.63 |
| BMI | 0.99 | 0.96, 1.03 | 0.74 |
| Obese (BMI>30) | 1.09 | 0.68, 1.76 | 0.71 |
| Diabetes | 1.33 | 0.76, 2.34 | 0.33 |
| Steroid Use | 1.89 | 0.71, 5.02 | 0.20 |
| Operative approach |  |  |  |
| Any MIS | 0.41 | 0.16, 1.03 | 0.058 |
| Unplanned open | 1.32 | 0.45, 3.83 | 0.62 |
| Neoadjuvant chemotherapy |  |  |  |
| Pre op systemic | 1.86 | 0.93, 3.70 | 0.077 |
| All other | 0.99 | 0.46, 2.14 | 0.99 |
| Number of mets resected |  |  |  |
| 3 – 4 | 0.73 | 0.35, 1.50 | 0.39 |
| 5 – 6 | 0.78 | 0.30, 2.06 | 0.62 |
| 7 – 8 | 1.32 | 0.53, 3.27 | 0.56 |
| > 8 | 2.16 | 1.18, 3.98 | **0.013*** |
| Size of lesion |  |  |  |
| 2-5 cm | 1.20 | 0.67, 2.14 | 0.53 |
| > 5cm | 0.88 | 0.45, 1.72 | 0.71 |
| Extent of resection |  |  |  |
| Total left lobectomy | 2.22 | 0.71, 6.89 | 0.17 |
| Trisegmentectomy | 0.68 | 0.13, 3.47 | 0.64 |
| Partial lobectomy | 1.82 | 0.77, 4.30 | 0.17 |
| CI, confidence interval; MIS, minimally invasive surgery; * indicates significance at p<0.05 | | | |
