## Supplemental Table 5 for "Role of ablation therapy in conjunction with surgical resection for neuroendocrine tumors (NETs): Risks and benefits of multimodality surgical treatment for NETs involving liver"

**Supplemental Table 5:** Univariate analysis of patient and procedure-related determinants of **any surgical site infection (n=117, 12.1%)** in patients undergoing resection of metastatic neuroendocrine tumors to the liver.

| Variable | Odds ratio | 95% CI | P value |
| --- | --- | --- | --- |
| Intraoperative ablation | 1.14 | 0.75, 1.72 | 0.54 |
| Age | 0.99 | 0.97, 1.00 | 0.13 |
| Sex | 0.98 | 0.67, 1.45 | 0.93 |
| Race group |  |  |  |
| Black/African American | 0.47 | 0.19, 1.20 | 0.12 |
| Other | 0.91 | 0.31, 2.64 | 0.86 |
| Not reported | 1.34 | 0.82, 2.17 | 0.24 |
| Ethnic group |  |  |  |
| Hispanic | 2.68 | 1.22, 5.89 | **0.014*** |
| Not reported | 1.38 | 0.83, 2.32 | 0.21 |
| Wound classification |  |  |  |
| I | 1.16 | 0.60, 2.25 | 0.65 |
| II | 1.64 | 0.59, 4.57 | 0.34 |
| III/IV | 1.06 | 0.12, 9.26 | 0.96 |
| BMI | 1.02 | 0.99, 1.05 | 0.18 |
| Obese (BMI>30) | 1.40 | 0.95, 2.07 | 0.092 |
| Diabetes | 1.31 | 0.81, 2.11 | 0.27 |
| Steroid Use | 1.48 | 0.60, 3.62 | 0.40 |
| Operative approach |  |  |  |
| Any MIS | 0.36 | 0.16, 0.80 | **0.012*** |
| Unplanned open | 1.06 | 0.40, 2.79 | 0.91 |
| Neoadjuvant chemotherapy |  |  |  |
| Pre op systemic | 1.20 | 0.63, 2.29 | 0.58 |
| All other | 0.66 | 0.32, 1.35 | 0.25 |
| Number of mets resected |  |  |  |
| 3 – 4 | 1.15 | 0.67, 1.96 | 0.61 |
| 5 – 6 | 1.12 | 0.54, 2.30 | 0.77 |
| 7 – 8 | 1.05 | 0.45, 2.42 | 0.92 |
| > 8 | 1.84 | 1.06, 3.19 | **0.029*** |
| Size of lesion |  |  |  |
| 2-5 cm | 1.42 | 0.86, 2.34 | 0.17 |
| > 5cm | 1.18 | 0.68, 2.06 | 0.56 |
| Extent of resection |  |  |  |
| Total left lobectomy | 1.12 | 0.44, 2.85 | 0.82 |
| Trisegmentectomy | 0.61 | 0.19, 2.00 | 0.41 |
| Partial lobectomy | 1.20 | 0.65, 2.23 | 0.56 |
| CI, confidence interval; MIS, minimally invasive surgery; * indicates significance at p<0.05 | | | |
